# Continuous theta-burst stimulation to a lesion network in cervical dystonia: a randomized, double-blind, sham-controlled, pilot trial

**DOI:** 10.64898/2026.09.10.26362783

**Authors:** Jordan Morrison-Ham, Ellen F. P. Younger, Elizabeth G. Ellis, Nicholas Parsons, Thomas Hart, Bridgette Speranza, Lillian Dipnall, Michael Do, Ruben Perellón-Alfonso, Pasquale Maria Pecoraro, Lazzaro di Biase, Kelly L. Bertram, Paul A. Agius, Francesca Morgante, Juho Joutsa, Aron T. Hill, Peter G. Enticott, Daniel T. Corp

**Author notes:** **Corresponding authors:** Jordan Morrison-Ham, School of Psychological Sciences & Turner Institute for Brain and Mental Health Monash University, Melbourne, Victoria, Australia, Daniel T. Corp, Cognitive Neuroscience Unit, School of Psychology, Deakin University, Burwood, Victoria, Australia.

## Abstract

**Background:** The mechanisms driving cervical dystonia remain unclear, limiting effective, non-invasive treatment development. Lesion network mapping previously localized a brain network derived from lesions causing cervical dystonia, with a network hub in the somatosensory cortex, a viable target for non-invasive brain stimulation.

**Objectives:** We conducted a randomized, sham-controlled, crossover trial administering continuous theta-burst stimulation (cTBS) to the S1 lesion network hub, testing whether modulating this network reduces CD symptoms.

**Methods:** Thirteen idiopathic cervical dystonia patients (57.85 ± 10.85 years) received two 10-session blocks of bilateral somatosensory cortex cTBS (active or sham; counterbalanced). The primary outcome was change in cervical dystonia symptoms, measured by the Toronto Western Spasmodic Torticollis Scale (TWSTRS). A wearable motion sensor objectively measured change in head and neck movement patterns. Motor evoked potentials and resting-state fMRI assessed cTBS-induced changes in corticospinal excitability and functional connectivity, respectively.

**Results:** Active cTBS did not significantly reduce cervical dystonia symptoms compared to sham (condition × timepoint interaction: *p* = .149). Range of motion significantly increased with active, versus sham, cTBS (*p* = .015). Corticospinal excitability decreased following sham (*p* = .042), but not active, cTBS. No significant connectivity changes were seen from the somatosensory cortex stimulation site after active cTBS; however, significant changes emerged from the globus pallidus interna.

**Conclusion:** This study prospectively tested the effects of neuromodulation on a lesion network mapping-derived target. Active cTBS did not significantly improve TWSTRS scores relative to sham. The intervention was well tolerated by all patients, and future studies could consider higher-dose or alternative protocols to this target.

## 1. Introduction

Cervical dystonia (CD) is characterised by excessive, involuntary contractions of the neck muscles.^1,2^ Whilst botulinum neurotoxin (BoNT) injections to the affected muscles can provide therapeutic relief^1^, up to one-third of CD patients report inadequate treatment response.^3^ In severe cases, deep brain stimulation (DBS) of the globus pallidus interna (GPi) or subthalamic nucleus can be effective^4,5^; yet, DBS is invasive and may cause unwanted side effects^4,6^. Transcranial magnetic stimulation (TMS) has been proposed as an alternative, non-invasive method to directly modulate these abnormal brain networks.^7^

To date, TMS has shown variable results in reducing CD symptoms.^8–12^ Clinical TMS protocols for CD are heterogeneous, particularly the therapeutic brain regions targeted.^13^ Although idiopathic dystonia is a network disorder involving neuroimaging abnormalities in multiple brain regions^14–16^, the correlational nature of many previous neuroimaging studies leaves it unknown whether these brain regions are driving the disorder, or are, for example, downstream changes or compensatory mechanisms.^17^ Thus, strong *a-priori* evidence regarding which regions might represent the most efficacious targets for TMS trials has been limited.

Lesion network mapping in CD has attempted to address this issue by localising a brain network functionally connected to lesions causing CD.^18,19^ Corp et al.^18^ demonstrated that whilst lesions causing CD were located throughout the brain, they were functionally connected to a common brain network. Notably, within this network, the cerebellum (positive connectivity) and somatosensory cortex (S1; negative connectivity) were functionally connected to all causal lesions, and this connectivity was specific compared to lesions causing other movement disorders.^18^ The clinical relevance of this lesion network was demonstrated in a cohort of patients who received GPi-DBS for idiopathic CD, with stronger functional connectivity from their GPi-DBS target location to the S1 network node showing better treatment outcomes.^18^

Based on this line of evidence, we previously investigated modulation of this S1 node prospectively with TMS.^20^ Here, positron emission tomography assessed the acute metabolic effect of a single continuous theta-burst stimulation (cTBS; a TMS protocol often regarded as inhibitory^21^) application to the lesion network mapping-derived S1 cluster. Results demonstrated abnormal local (S1) metabolic changes, and remote network effects (brainstem) in CD patients.^20^ Whilst these results demonstrate target engagement, whether neuromodulation of this brain region can reduce CD symptoms remains untested. More broadly, although lesion network mapping has localised brain networks in over 40 neurological symptoms^22–26^, to our knowledge, no study has targeted a brain network derived from lesion network mapping. Thus, the utility of this method to localise targets for clinical neuromodulation remains unknown.

Therefore, we aimed to test whether a multi-day cTBS protocol to the S1 cluster localised by lesion network mapping^18^ would reduce CD symptoms. We conducted a randomized, double-blind, sham-controlled, crossover pilot trial using cTBS in idiopathic CD patients. The primary outcome was changes in CD experience measured by the Toronto Western Spasmodic Torticollis Rating Scale (TWSTRS).

## 2. Methods

### 2.1 Patients

Fourteen individuals diagnosed with idiopathic CD enrolled in the study (July 2022 to November 2024). One patient withdrew after three sessions for personal reasons; thus, 13 patients contributed to the primary outcome analysis. All patients provided written informed consent and were screened for MRI and TMS contraindications before enrolment.

Inclusion criteria were: 1) aged 18-85 years; 2) an idiopathic CD diagnosis. Exclusion criteria were: 1) current or past history of a major psychiatric disorder (e.g., schizophrenia) or neurological condition other than CD; 2) prior diagnosis of brain injury or lesion; 3) metal implants in the skull; 4) history of seizure; 5) current or past history of substance abuse or dependence; and 6) pregnancy. Patients prescribed BoNT injections observed a minimum 6-week washout period prior to beginning a cTBS block, to minimize the effects of BoNT on primary outcomes and ensure patients were able to participate without experiencing treatment delays.

### 2.2 Study Design

We implemented a randomized, sham controlled, double-blind, crossover design. Patients received two, 2-week blocks of cTBS (one active and one sham block) to the S1 bilaterally. Each block consisted of 10 consecutive weekday cTBS sessions. There was a minimum two-week washout period between blocks to minimise carry-over effects. Patients and assessors administering the clinical outcome measures were blinded to allocation, while TMS operators were not. See Figure 1 for experimental protocol, Supplementary Methods for randomization and sample size calculations.

**Figure 1.**
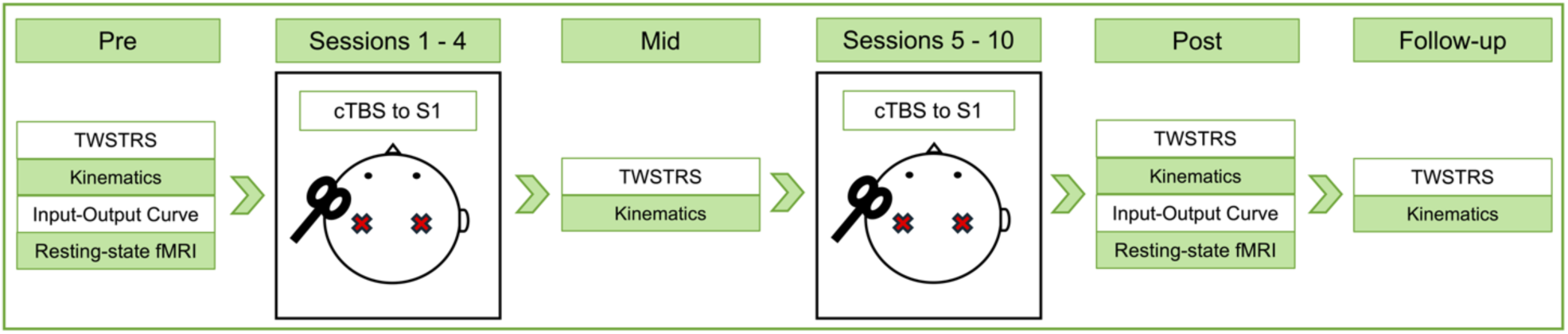
Experimental design. Each block of stimulation, regardless of condition (i.e., active or sham cTBS) followed the same procedures. Before the application of cTBS (Pre), patients received MRI scans and completed the TWSTRS, movement kinematics, proprioception and input-output curve measures. Prior to the application of cTBS in the fifth session, patients again completed the TWSTRS, kinematics and proprioception measures (Mid). At least 90 minutes after the application of cTBS in session 10, TWSTRS, kinematics, proprioception, input-output curves, and MRI scans were conducted (Post). Patients returned approximately 2 weeks later to complete the TWSTRS, movement kinematics and proprioception measures (Follow-up). This protocol was then repeated for the second block of stimulation.

Before the application of cTBS (Pre), patients received MRI scans and completed the TWSTRS, movement kinematics and proprioception, and single-pulse TMS input-output curve measures. Prior to the application of cTBS in the fifth session, patients again completed the TWSTRS, kinematics and proprioception measures (Mid). At least 90 minutes after the application of cTBS in session 10, TWSTRS, kinematics and proprioception, input-output curves, and MRI scans were conducted (Post). Patients returned approximately 2 weeks later to complete the TWSTRS, movement kinematics and proprioception measures (Follow-up). Pre and Mid outcomes were assessed before stimulation application in sessions 1 and 5, and post-intervention outcomes were assessed 90 minutes after stimulation in session 10 to avoid acute brain stimulation effects.^27^ MRI scans were conducted maximum three days before and after each stimulation block (Pre and Post only). After the final follow-up session, once all outcomes were collected, patients were asked to guess the order of stimulation (active/sham) they received.

Study procedures were approved by the Deakin University Human Research Ethics Committee (DUHREC 2021-136) and were conducted in accordance with the Declaration of Helsinki.^28^ The study was prospectively registered on the Australian New Zealand Clinical Trials Registry in April 2021 (ACTRN12621000417886). See Supplementary Material, Figure S1 for CONSORT^29^ flow diagram and checklist.

### 2.3 Primary Outcome

The primary outcome measure was the TWSTRS total score (range: 0-85), with higher scores indicating more severe symptoms.^30^ TWSTRS data was collected at all timepoints across each block (i.e., Pre, Mid, Post, Follow-up; Figure 1).

### 2.4 Secondary Outcomes

#### 2.4.1 TWSTRS Subscales

The three TWSTRS subscales of severity (0-35), disability (0-30), and pain (0-20) were each analysed secondarily.

#### 2.4.2 Movement Kinematics and Proprioception

Patients’ head and neck movement kinematics were captured using an XSENS Dot wearable sensor, placed on the inion with a headband. Starting with their head as close to a central position as possible, patients were asked to: 1) turn their head as far to the left and as far to the right as comfortably possible, while keeping shoulders and lower body still, before returning to their central position; 2) look upward and then downward as far as comfortably possible; 3) tilt their ear to their shoulder on the left then right as far as comfortably possible. All movements were repeated twice. The XSENS device measured acceleration and angular velocity of the movements on pitch (x), roll (y), and yaw (z) axes.

In addition, patients completed a proprioception task (based on Avanzino et al.^31^) whilst wearing the sensor, motivated by suggestions that CD may involve abnormal processing of proprioceptive information.^18,32–34^ Patients were asked to close their eyes, while their head was passively displaced by an investigator to a position on one of four axes (left, right, up, down). This ‘passive’ position was held for 5 seconds, before the investigator returned their head back to their central position. In the active trial, the patient was asked to replicate this head position in space and hold this for 3 seconds, before returning back to their central position.

These tasks were completed at all timepoints across each block (Figure 1).

#### 2.4.3 Input-Output Curves

Input-output curve data was collected in the first session (before cTBS; Pre) and the last session (90 minutes after cTBS; Post) of both stimulation blocks, using single TMS pulses over the left and right first dorsal interosseous (FDI) muscle hotspots within the motor cortex (M1). Blocks of ten stimuli were delivered in pseudorandom order (50%, 40%, 60%, 70% of maximum stimulator output [MSO]), at random 5-8s intervals to avoid stimulus anticipation.^35^

#### 2.4.4 Magnetic Resonance Imaging

MRIs were conducted before (Pre) and after (Post) each cTBS block (Figure 1). Data were acquired using a 3T Siemens Skyra scanner with a 32-channel head coil. At the baseline MRI scan, a T1-structural image (MPRAGE) was acquired for all patients for neuronavigation purposes with the following parameters: TA = 5:08 minutes, TR = 1860.0 ms, TE = 2.18 ms, TI = 1000 ms, voxel size = 0.8 mm x 0.8 mm x 0.8 mm, flip angle = 8°, field of view = 256 mm x 256 mm, 208 sagittal slices. For all MRI sessions patients underwent a 16 minute resting-state fMRI sequence, comprising two resting-state fMRI runs with reverse phase encoding directions (A-P, P-A), with the following parameters: TA = 8 minutes, TR = 1486 ms, TE = 36.60 ms, voxel size = 2.5 mm x 2.5 mm x 2.5 mm, flip angle = 60°, field of view = 250 mm x 250 mm, multiband acceleration factor = 4.

### 2.5 Electromyography

Electromyography, motor hotspot identification, and resting motor threshold were performed according to established guidelines.^36,37^ Participants were seated in a comfortable chair with their hands at rest. Bipolar surface electromyography (EMG) was recorded from the left and right FDI muscles using 20mm Ag/AgCI surface electrodes placed over the muscle belly and corresponding tendon, with a reference electrode on the right third knuckle. EMG signals were collected using a PowerLab 4/35 data acquisition device (ADInstruments, USA), amplified (gain × 1000), band-pass filtered (1–2000 Hz), and sampled at 4 kHz as per IFCN guidelines.^37^ EMG activity was recorded from 100ms before to 100ms after each TMS stimulus. The root mean square of the EMG amplitude from 100 to 0ms prior to the TMS pulse was calculated to analyse resting muscle activity. EMG signals were recorded by a computer using Labchart version 8 software (AD Instruments, USA).

### 2.6 Single Pulse TMS

Patients underwent daily screening to monitor known factors that impact TMS safety (e.g., daily sleep hours, alcohol consumption levels, or substantial changes to nicotine or caffeine intake^38,39^). Motor hotspot and input-output response curves were measured using single-pulse TMS, delivered via a Magstim 2002 unit with a 70mm figure-of-eight handheld coil (Magstim Company Ltd, UK). This machine was used as per evidence that monophasic pulses are more appropriate than biphasic pulses to measure the after-effects of theta-burst stimulation.^40^ The coil was held tangentially to the scalp with the handle posterior to the head, positioned approximately 45° to the midline. The motor hotspot was determined in the first session of the first block for both hemispheres, initially by orientating the coil to the participant’s M1 “hand knob” representation via the use of neuronavigation. The coil was then systematically moved around multiple locations within a search grid spanning approximately 2cm in all directions, collecting at least 5 motor evoked potentials (MEPs) at each location, to find the site giving the largest and most consistent MEP.^36^ This site was then defined as the ‘motor hotspot’ and recorded in the neuronavigation program.

### 2.7 Neuronavigation and S1 Targeting

Neuronavigation was used in conjunction with structural MRI scans to localize the S1 intervention target for each participant. T1-weighted MRI images were imported into Brainsight (version 2.4.9; Rogue Research Inc.), where MRI-to-head co-registration was performed. The co-registered brain was adjusted according to the participants anterior-posterior commissure line, transforming the brain into Montreal Neurological Institute (MNI) space. This co-registration method was independently verified for each participant with a custom-made T1 MRI preprocessing script to ensure the same brain region was localised. The anterior-posterior commissure line method was preferred as it resulted in a more realistic skin reconstruction, allowing for better coil position and trajectory planning. Head registration landmarks were placed on the nasion and bilateral tragi on all participants. The cTBS targets for stimulation were positioned bilaterally in the S1 at the following MNI coordinates: left MNI x = −45, y = −30, z = 58, right MNI x = 45, y = −30, z = 58. These targets were selected based on lesion network mapping by Corp et al.^18^ which demonstrated that 25/25 lesions causing CD were connected to a cluster of voxels in the right S1 (centre of gravity: x = 45, y = −30, z = 58; previously used as the stimulation target in the preliminary metabolic response to cTBS study by Kokkonen et al.^20^). Bilateral stimulation was chosen because: 1) both S1 hemispheres were part of the final lesion network and similarly connected to causal lesions^18^; 2) to maximise the possible clinical effects of cTBS; 3) to avoid possible hemispheric imbalances in excitability as a result of the cTBS.

We used an individualised, sulcus-aligned approach to optimise the TMS coil placement according to each patient’s brain anatomy. First, a 3D model of the participant’s brain, computed from their T1 scan, was created in Brainsight. The MNI coordinates for the bilateral S1 treatment sites were entered and coil-shaped markers were placed on the 3D model at the target site. Next, the coil twist was individualised to align the coil marker perpendicular to the central and post-central sulci in the brain, at the point of the specific cTBS stimulation site for each participant. This was done in an attempt to align the induced electric field in the brain parallel to the assumed direction of neurons within the S1 target region, which has been shown to result in greater neuronal activation.^41^ To ensure the coil lay flat on the head, minimising the distance between the coil and the cortex, the tilt of the coil marker was then optimised to the participants head model.

### 2.8 Resting Motor Threshold Assessment

To determine the threshold at which to deliver the intervention, each participant’s resting motor threshold (RMT) was identified using the TMS Motor Threshold Assessment Tool (MTAT 2.0; with the option for assessment without a priori information selected)^42^, before cTBS in the first session of the first block. An MEP for RMT assessment was defined as a recognisable peak-to-peak deflection 50 microvolts in amplitude applied over the motor hotspot, at the appropriate latency of a hand muscle MEP (approximately 20-25 milliseconds after the TMS pulse^37^). RMT assessment and was delivered using a Magventure Magpro Rapid R30 (Magventure, Farum, Denmark), with a figure-of-eight 75mm cooled coil (MCF-B65).

### 2.9 Intervention: Continuous Theta-Burst Stimulation

As in our recent study^20^, cTBS was selected for its short stimulation time and prior evidence of hyperactivity within the S1 of CD patients.^18,39,43^ cTBS was delivered using a Magventure Magpro Rapid R30 (Magventure, Farum, Denmark), with a figure-of-eight 75mm cooled coil (MCF-B65), applied at 70% of a patient’s resting motor threshold to the bilateral S1 (left MNI x = −45, y = −30, z = 58, right MNI x = 45, y = −30, z = 58; based on lesion network mapping results from Corp et al.^18^). cTBS consisted of three-pulse bursts at 50Hz repeated every 200ms (5Hz) for 40 seconds (600 pulses total^21^). The sham cTBS block was identical but delivered using a figure-of-eight 75mm cooled placebo coil (MC-P-B65) which produces a similar sensation and sound to the active coil but with a reduced magnetic field.^44^

All TMS procedures were conducted bilaterally, with application first over the left then right hemisphere. For the cTBS intervention, an interval of approximately one minute separated each hemisphere.

### 2.10 Statistical Analysis

Linear mixed effects models using restricted maximum likelihood estimation (REML) were conducted for the TWSTRS, movement kinematic outcomes, proprioception, and input-output curves, and were performed in R Studio (version 2025.05.1.513^45^) using packages lme4 (version 1.1.37^46^), lmerTest (version 3.1.3^47^) and emmeans (version 1.11.1^48^). Degrees of freedom for fixed effects were estimated using Satterthwaite approximation^47^, with analyses considered significant if *p* < .05. Where significant interactions were found, Bonferroni corrected post-hoc simple effects were analyzed.

#### 2.10.1 TWSTRS Scores

To assess changes in CD symptoms (TWSTRS total score), timepoint (levels: pre, mid, post, and follow-up) and cTBS condition (levels: active and sham) were included as fixed effects, and patient ID entered as a random intercept to account for the data clustered within patients. Separate models were conducted for the TWSTRS total score and subscales.

#### 2.10.2 Movement Kinematics and Proprioception Task Outcomes

Three kinematic outcome measures were analysed for each movement kinematics task: 1) range of motion; 2) peak angular velocity; and 3) ‘movement smoothness’, where higher values indicated greater smoothness. Timepoint and cTBS condition were included as fixed effects, and patient ID as a random intercept. Separate models were run for each outcome measure, on each movement axis (i.e., up-down, left-right, tilt). For the proprioception task, the relative angular error between the active and passive head positions was analysed using a similar mixed-effects model. See Supplementary Methods 1.3 for further detail on outcome extraction, calculation, and analysis.

#### 2.10.3 Input-Output Curve

For single-pulse TMS, changes in MEP amplitudes due to cTBS condition were analyzed. The mean MEP amplitude value was calculated for each block of pulses and used as the dependent variable – no MEP data were deleted or transformed.^49^ Timepoint, cTBS condition, and stimulator intensity (levels: 40%, 50%, 60%, 70%) were entered as fixed effects, with patient ID entered as a random intercept to preserve nesting of data within patients. The root mean square of the EMG trace 100-0ms before the TMS pulse was entered as a covariate to control for involuntary muscle activity. Models were conducted for right and left hemispheres separately.

To test for an association between corticospinal excitability and symptom severity, correlations were performed between raw changes in TWSTRS total score and MEP amplitude. Analyses were run separately for each stimulator intensity in each condition (active or sham).

#### 2.10.4 fMRI Preprocessing and Analysis

fMRI data preprocessing and analysis was performed using standardised pipelines with CONN (version 22^50^) and SPM12 (https://fil.ion.ucl.ac.uk/spm/) toolboxes in MATLAB (version R2024b; Mathworks, USA); full details are presented in the supplementary methods (section 1.4). Briefly, to test for stimulation induced changes in functional connectivity, seed to whole brain connectivity analyses were conducted. Six seeds from two brain regions were defined *a priori.* The primary seeds of interest were those at the centre of gravity of the bilateral and unilateral S1 cTBS stimulation sites (MNI x = ±45, y = −30, z = 58; 4mm seeds). An *a priori* exploratory analysis was also run from 4mm seeds generated at the bilateral and unilateral GPi (derived from the optimal site for deep brain stimulation in CD^51^; x = ±19.4, y = −10.1, z = −5.9 mm). TWSTRS total score was added as a covariate to test for associations between connectivity and changes in symptom scores. For clusters found in each analysis, threshold-free cluster enhancement (TFCE) was applied using 5000 permutations, with results considered significant at a family-wise error (FWE) corrected threshold of *p* < .05^52,53^.

Because not all patients received fMRI scans in both active and sham conditions, analyses were restricted to comparing individual cTBS conditions across time (main effect of time within the active and sham cTBS condition separately, rather than the interaction of timepoint and condition). One patient was removed from analyses due to significant motion, leaving *n* = 10 for both active and sham cTBS analyses. The 10 patients differed between conditions (Table 1).

**Table 1.** Patient descriptive characteristics.

|  | Major symptom | Sex | Time since diagnosis (years) | Additional symptoms | Mean BoNT schedule (weeks) | Time between TMS blocks (weeks) | TMS doses (active block) | TMS doses (sham block) | MRI scans received (active block) | MRI scans received (sham block) |
| --- | --- | --- | --- | --- | --- | --- | --- | --- | --- | --- |
| 1 | Torticollis | F | 25 | None | 12 | 12 | 10 | 9 | 2 | 0 |
| 2 | Torticollis/Laterocollis | F | >1 | Leg dystonia | 12 | 3 | 5 | 5 | 2 | 0 |
| 3 | Torticollis | F | 4 | None | 12 | 2 | 9 | 9 | 2 | 0 |
| 4 | - | F | 5 | None | 20 | 3 | 10 | 10 | 1 | 2 |
| 5 | Retrocollis | F | 2 | Blepharospasm, oromandibular dystonia | 12 | 4 | 10 | 10 | 2 | 2 |
| 6 | Torticollis/Laterocollis | F | 17 | None | - | 2 | 10 | 10 | 2 | 2 |
| 7 | Torticollis/Laterocollis | M | 3 | None | - | 2 | 9 | 9 | 2 | 0 |
| 8 | Torticollis/Anterocollis | F | 2 | Facial spasm | 12 | 2 | 10 | 10 | 2 | 2 |
| 9 | Torticollis/Retrocollis | F | 10 | None | 12 | 8 | 9 | 10 | 2 | 2 |
| 10 | Torticollis | F | >1 | None | 12 | 2 | 10 | 10 | 2 | 2 |
| 11 | Anterocollis | F | >1 | None | 12 | 2 | 10 | 10 | 1 | 0 |
| 12 | Torticollis | M | >1 | None | 12 | 2 | 10 | 10 | 2 | 2 |
| 13 | Torticollis | F | 4 | None | 12 | 2 | 10 | 10 | 2 | 2 |

*Note.* F = Female; M = Male; BoNT = botulinum neurotoxin; TMS = transcranial magnetic stimulation; MRI = magnetic resonance imaging.

## 3. Results

### 3.1 Patient Demographics

Patient mean age was 57.85 (*SD* = 10.85), with time since CD diagnosis ranging from one to 25 years (Table 1). One patient had no major symptom pattern, scoring 0 on the TWSTRS at all study timepoints but one, yet were retained in analyses as they had a diagnosis of CD and met all inclusion criteria. Eleven patients were actively receiving BoNT injections prior to trial enrolment (mean time since injection = 8 ± 2.32 weeks, range = 6-13 weeks). The two remaining patients had stopped receiving injections and did not intend to continue. Three patients maintained a low dose of benzodiazepines, with patients instructed to maintain their normal medication regimen during trial participation.

Standardised post-stimulation questionnaires administered at the end of each session indicated that patients tolerated cTBS well, with only minor adverse events reported (headache *n* = 2; sensitivity to light *n* = 1; fatigue *n* = 1; scalp discomfort *n* = 1). Twelve of the 13 patients completed ≥ 9 cTBS sessions in both active and sham conditions, while one patient completed 5 sessions for both conditions (missingness all due to unrelated illness or unavailability). These numbers were balanced across conditions (Table 1). Ten of 13 patients correctly guessed the order of stimulation they received (reasons for their guesses are provided in Table S2).

### 3.3 Primary Outcome: TWSTRS total score

Analyses demonstrated no significant time × condition interaction (*F*(3, 82) = 1.82, *p* = .149; Figure 2A). There was a significant main effect of timepoint (*F*(3, 82) = 3.12, *p* = .030), with *post-hoc* pairwise comparisons indicating that this effect was driven by significant reductions in TWSTRS scores from the Pre to Post timepoint, across the active and sham conditions (estimate = −3.30, 95% CI: −0.56 – −6.04, *p_corr_* = .013).

**Figure 2.**
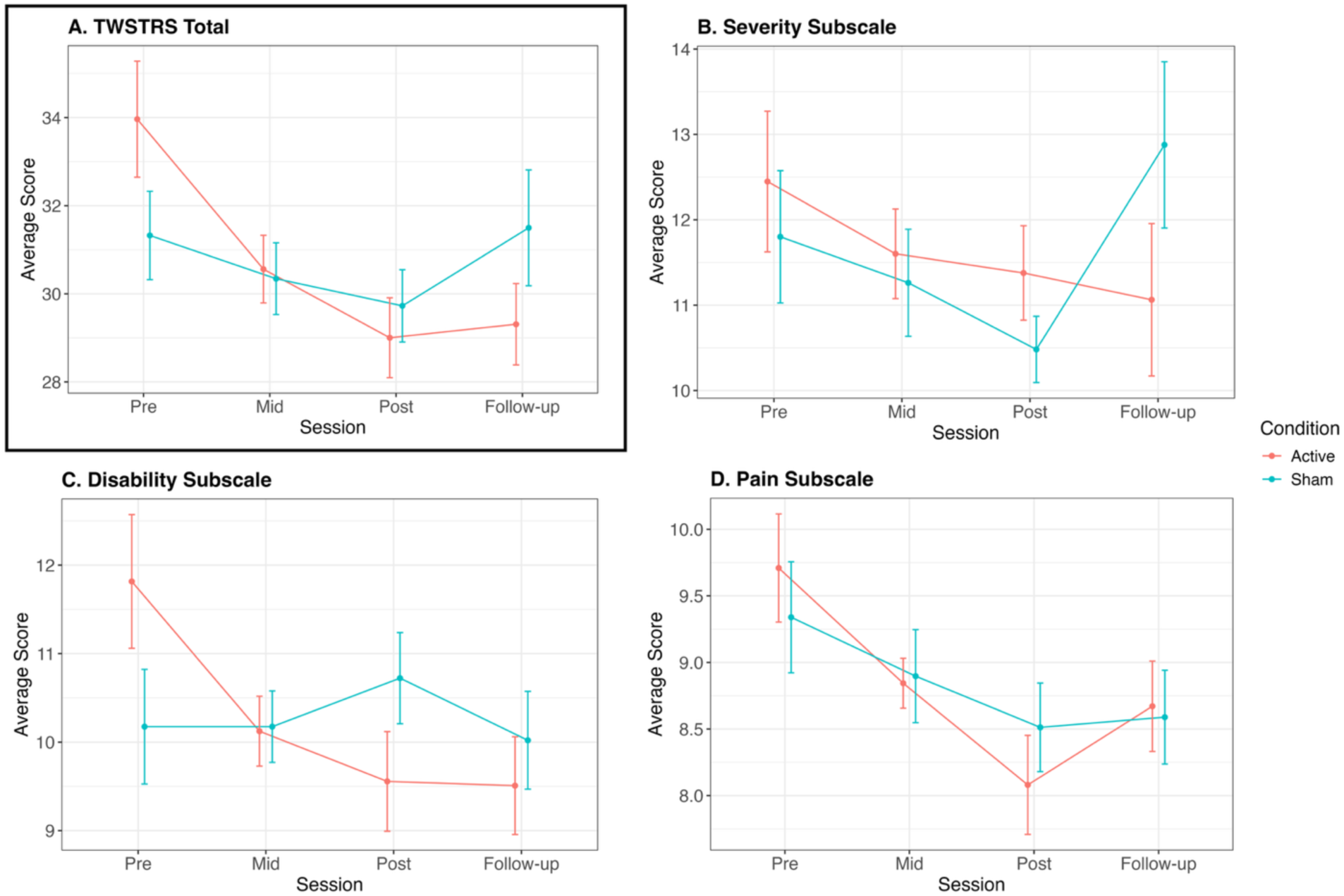
Average TWSTRS total (A) and subscale scores (B-D) across timepoints, by cTBS condition. Mean values and standard errors are presented. 13 CD patients were included in TWSTRS linear mixed effects models, with no significant interaction effects present.

Cohen’s *d* effect sizes between each of the timepoints were as follows: Active cTBS Pre – Mid = 0.19; Pre – Post = 0.20; Pre – Follow-up = 0.27; Sham cTBS Pre – Mid = 0.05; Pre – Post = –0.01; Pre – Follow-up = –0.01.

### 3.4 Secondary Outcome: TWSTRS subscales

There were no significant interaction effects for any of the TWSTRS subscales (severity: *p* = .259; disability: *p* = .098; pain: *p* = .706; Figure 2B-D). See Supplementary Results 2.1 for main effects.

### 3.5 Secondary Outcome: Movement Kinematics and Proprioception

There was a significant two-way interaction between cTBS condition and timepoint for range of motion on the left-right axis (*F*(3, 66.12) = 3.74, *p* = .015; Figure 3Aii). As there was a significant difference in the Pre timepoint scores between conditions (estimate = −8.24, 95% CI: −15.41 – −1.07, *p* = .025), *post-hoc* tests investigated simple interaction effects (i.e., the difference between Pre and all other timepoints across conditions). A significantly greater increase in range of motion was seen from Pre to Follow-up for the active, compared to sham, condition (estimate = −17.27, 95% CI: −30.40 − −4.12, *p_corr_* = .006).

**Figure 3.**
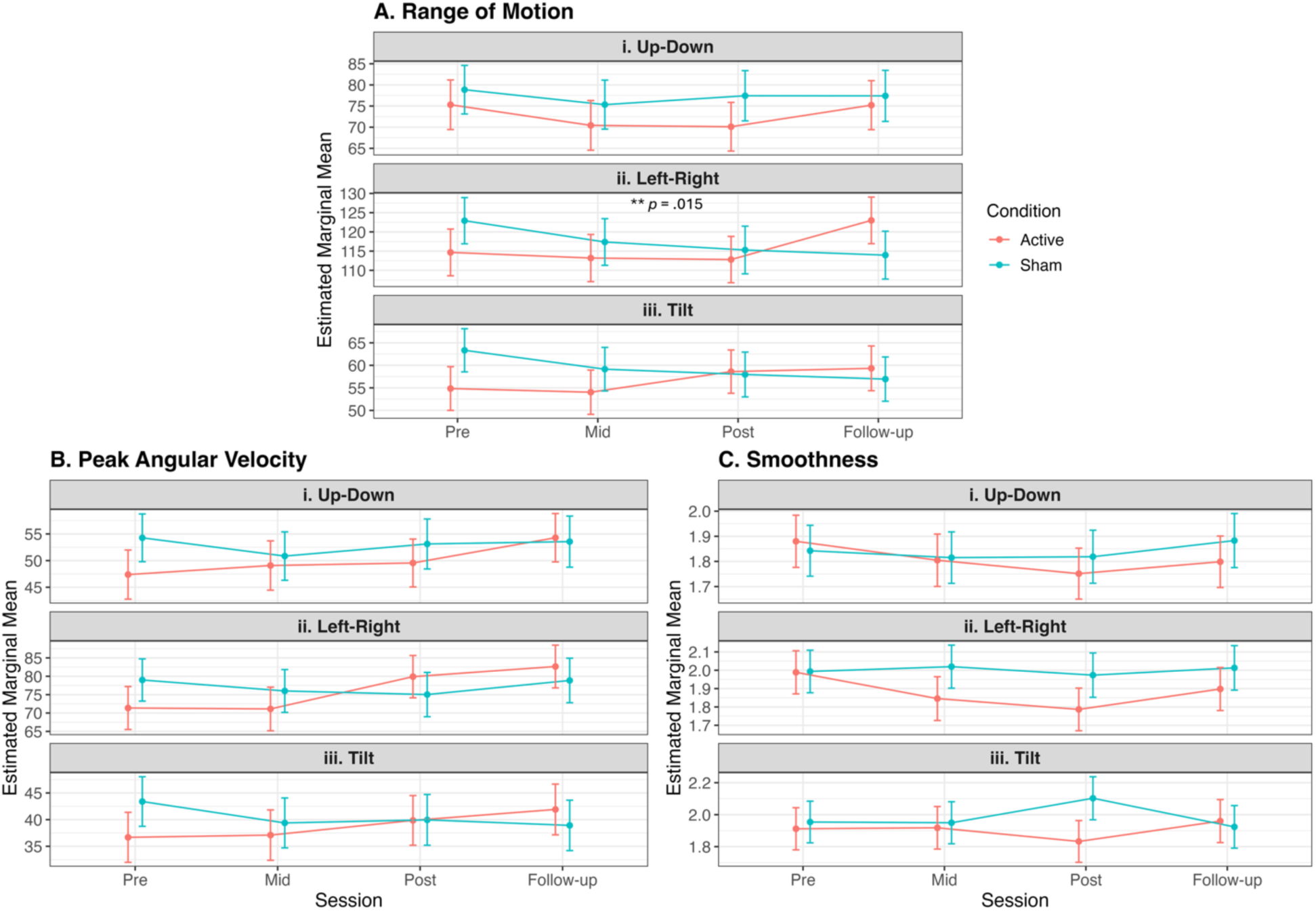
Model-predicted marginal means for each kinematic outcome and axis, across timepoints and conditions. (A) Range of motion. Linear mixed effects models demonstrated a significant increase in the left-right axis (Aii) range of motion for the active group compared to the sham (*p* = .015). (B) Peak angular velocity. (C) Smoothness. Error bars indicate standard error. 12 CD patients were included in the mixed effects models

**Figure 4.**
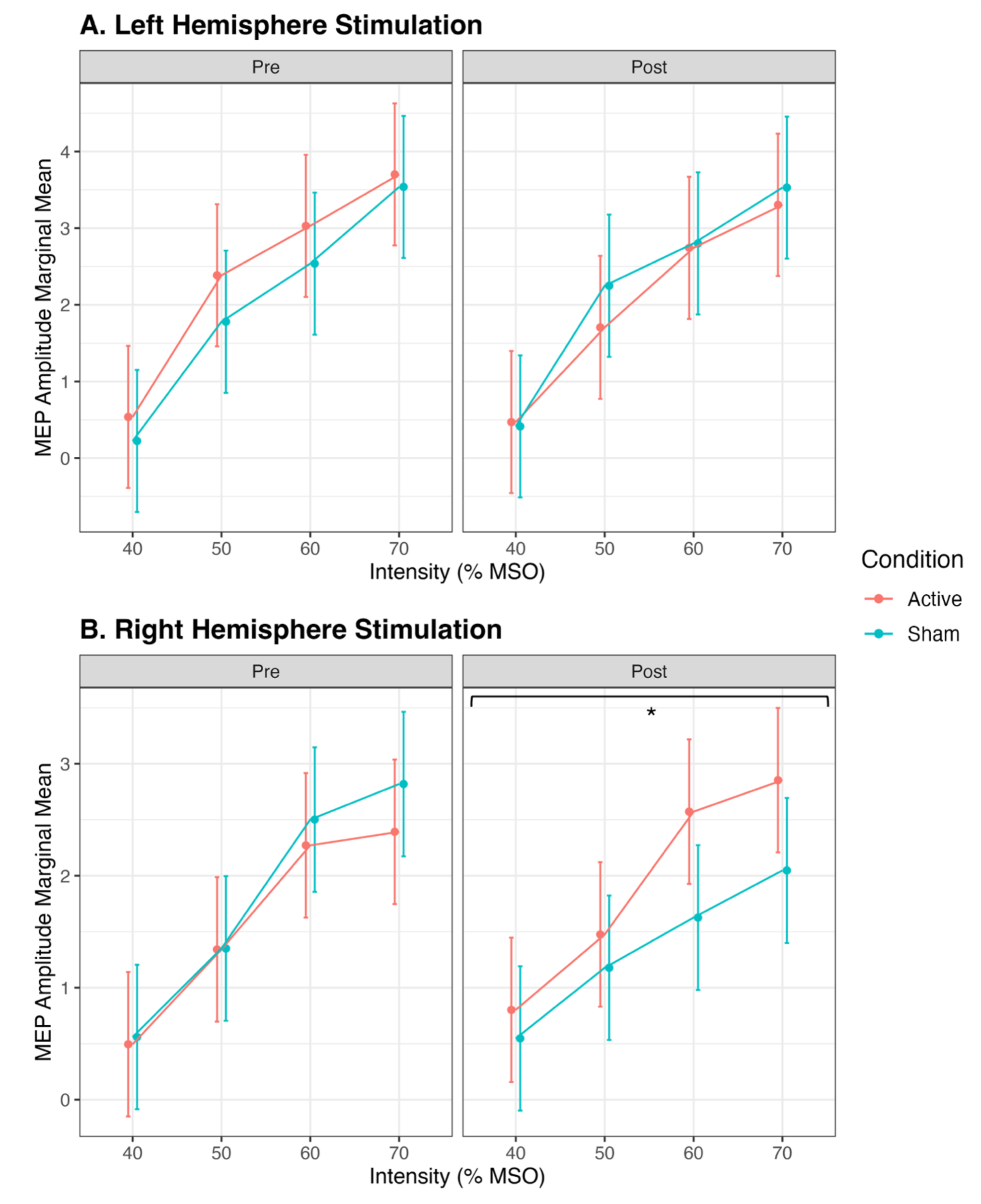
Model-predicted MEP amplitudes (marginal means) across stimulation intensities and conditions, controlling for pre-stimulation EMG. (A) Left hemisphere. (B) Right hemisphere, with mixed effect models demonstrating a significant reduction in MEP amplitude from Pre to Post in the sham condition (*p* = .042). Error bars indicate standard error. 12 CD patients were included in IO curve analyses.

There was a trend towards significance for a two-way interaction on the tilt axis of the peak angular velocity metric (*F*(3, 65.07) = 2.66, *p* = .056; Figure 3Biii), and the tilt axis of the smoothness metric (*F*(3, 65.00) = 2.58, *p* = .061; Figure 3Ciii). No proprioception outcomes were significant (Supplementary Results 2.2).

### 3.6 Secondary Outcome: Input-Output Curve

Left hemisphere analyses indicated no significant two- or three-way interaction effects (timepoint × condition × stimulus intensity: *p* = .907; timepoint × condition: *p* = .980; Figure 3A).

There was no significant three-way interaction (*p* = .552) for right hemisphere stimulation, however, there was a significant two-way (timepoint × condition) interaction (*F*(1, 163.81) = 6.06, *p* = .014). *Post-hoc* pairwise comparisons demonstrated that this interaction was primarily driven by a significant reduction in MEP amplitude (across intensities) from Pre to Post in the sham cTBS group (estimate = −0.46, 95% CI: −0.90 – −0.02, *p* = .042; Figure 3B), while the increase in MEP amplitude from Pre to Post in the active cTBS group was not significant (estimate = 0.30, 95% CI: −0.13 – 0.73, *p* = .173).

There were no significant associations between change in MEP amplitude and TWSTRS scores from pre to post cTBS, for active or sham conditions (Supplementary Figure S2).

### 3.7 Secondary Outcome: fMRI Analysis

There were no significant changes in functional connectivity from the S1 primary seeds of interest in either active or sham conditions for the whole brain analysis, nor when constrained to the lesion network from Corp, et al.^18^

In the exploratory analysis examining connectivity from the GPi, there was a significant decrease in positive functional connectivity after active cTBS (from Pre to Post), from the right GPi to the right temporal pole (whole brain P_FWE_ = .012; Supplementary Figure S3a).

There was also a significant negative association between the change in functional connectivity from the right GPi to the right motor cortex, and the change in TWSTRS total score from the Pre to the Post timepoints after active cTBS (whole brain P_FWE_ = .038; Supplementary Figure S3b). Specifically, a shift towards more positive connectivity between these two brain regions after active cTBS was associated with a reduction in TWSTRS score.

No other analysis survived multiple comparisons correction. See Supplementary Table S2 for significant uncorrected findings.

## 4. Discussion

We conducted a randomized, sham-controlled, crossover pilot trial of 10 days of cTBS to the S1 in idiopathic CD patients. To the best of our knowledge this is the first clinical trial to prospectively test the effects of neuromodulation of a brain network derived from lesion network mapping.^18^ There were no significant changes in the primary outcome (TWSTRS) after active cTBS, when compared to sham stimulation. However, active cTBS significantly increased patient range of motion across a left-right movement axis and resulted in changed in neurophysiological outcomes as measured by single-pulse TMS and rs-fMRI. As such, the results do not provide support for therapeutic efficacy of cTBS to the S1 in CD, yet these results warrant further investigation of this question. Future studies may consider higher-dose TMS protocols to this CD network.

### Effects of cTBS to the lesion network on symptoms of cervical dystonia

While TWSTRS scores did not show a significant effect of active cTBS, there was a significant increase in patient range of motion for active vs sham cTBS. This could represent subtle changes in patient symptoms that were not captured by the clinical scale. However, this could also represent placebo effects, with most patients correctly guessing treatment condition.

Considering reasons for the lack of significance on the primary outcome, we did not reach our recruitment goal of 20 patients, which may have resulted in a lack of statistical power. Moreover, the cTBS dose (once per day for 10 days) may have been too low to drive meaningful clinical improvements. Accelerated TMS protocols have recently shown improved outcomes in depression, whilst increasing feasibility and practicality.^54,55^ The application of accelerated cTBS in neurological disorders, in particular visuospatial neglect, suggests that increased cTBS doses may be necessary to induce clinically meaningful and durable symptom improvement.^56,57^ Thus, future work could consider accelerated cTBS protocols to optimise therapeutic efficacy in CD.

### Higher corticospinal excitability after active cTBS compared to sham cTBS

Evidence of increased cortical excitability and S1 hyperactivity in dystonia^58–62^ motivated the use of a cTBS in this study, a technique typically thought to reduce cortical excitability at the stimulated site.^21^ Therefore, we expected that cTBS would also result in reduced corticospinal excitability. Contrary to this, MEP amplitudes decreased following sham, but not active, cTBS. In our prior study measuring metabolic response to a single cTBS session to the same S1 network node^20^ there was also significantly higher metabolic activity at the stimulation site for CD patients who received active cTBS compared to sham. Although methods differed, and MEP amplitudes were not measured in that study, it is interesting to note that both studies observed a general pattern of increased excitability or activity in the sensorimotor cortex in CD patients, when the opposite effect would be expected. It is now appreciated that responses to theta-burst stimulation are far more complex than simply increasing or decreasing neural activity within the stimulated area^63,64^, and it cannot yet be said which pattern of response is desired after therapeutic TMS in CD patients. Future studies could consider comparing multi-day continuous and intermittent theta-burst stimulation, to investigate possible dissociations in neurophysiological responses and clinical improvement.

### Changes in resting-state functional connectivity

Active cTBS produced no significant changes in functional connectivity from S1 stimulation sites; however, significant changes were observed from the GPi after active, but not sham, cTBS. These findings suggest that cTBS applied to the S1 network node, identified via lesion network mapping, may engage a similar brain network to that modulated by GPi DBS. This agrees with prior research demonstrating changes in functional connectivity to the sensorimotor cortices after GPi DBS, that were associated with improvements in clinical symptoms.^62,65^ However, even with a repeated measures design, a sample size of 10 patients is small for rs-fMRI analyses, and these effects should be replicated in a larger sample. Studies using similar TMS therapeutic protocols in CD would allow for pooled analyses to better detect cardinal patterns of functional connectivity changes that are associated with therapeutic response.

### Limitations

This study had several limitations. First, ten of 13 patients correctly identified the order of cTBS stimulation they received (Supplementary Table S1). Therefore, we cannot exclude the possibility of placebo effects. We elected to employ a crossover design because of concerns about patient recruitment during restrictive COVID-19 lockdowns in Melbourne, Australia, yet parallel designs may be preferable to maintain blinding. Moreover, the sample size was relatively small, and we did not reach our target recruitment goal. Therefore, we may have lacked statistical power to demonstrate significant effects. Further, neurophysiological responses measured using single-pulse TMS and fMRI are variable, thus our observed effects will need to be replicated in larger CD cohorts. Finally, DBS in dystonia can require months for symptom improvement to be demonstrated^66,67^, and our short two-week follow-up period may have failed to capture longer term changes.

## Conclusions

This prospective pilot clinical trial tested the effects of neuromodulation of a lesion network mapping-derived brain network in CD. Relative to sham, active cTBS did not significantly reduce CD symptoms on the TWSTRS scale (primary outcome). Significant effects were observed in secondary outcomes, including changes in head movement range of motion, and neurophysiological outcomes. Future studies could investigate the effect of neuromodulation to this lesion network mapping derived target using accelerated or alternative TMS protocols.

## Supporting information

All supplementaries

## Author Roles

(1) Research Project: A. Conception, B. Organization, C. Execution; (2) Statistical Analysis: A. Design, B. Execution, C. Review and Critique; (3) Manuscript Preparation: A. Writing of the First Draft, B. Review and Critique; (4) Other: A. Funding Acquisition.

**JM-H:** 1A, 1B, 1C, 2A, 2B, 2C, 3A, 4A.

**EFPY:** 1B, 1C, 2C, 3B.

**EGE:** 1B, 1C, 2C, 3B.

**NP:** 2A, 2B, 2C, 3B.

**TH:** 2A, 2B, 2C, 3B.

**BS:** 1C, 3B.

**LD:** 1C, 3B.

**MD:** 1C, 3B.

**RP-A:** 1B, 2C, 3B.

**PMP:** 2B, 2C, 3B.

**LdB:** 2B, 2C, 3B.

**KLB:** 1A, 1B, 3B.

**PAG:** 2C, 3B.

**FM:** 2C, 3B.

**JJ:** 1A, 2A, 2C, 3B.

**ATH:** 1C, 2A, 2C, 3B.

**PGE:** 1A, 1B, 2C, 3B.

**DTC:** 1A, 1B, 1C, 2A, 2B, 2C, 3B, 4A.

## Data Availability

The deidentified data that support the findings of this study are available at the Big NIBS data repository (bignibsdata.com) and OpenNeuro (https://openneuro.org/datasets/ds008174).

## Acknowledgments

The authors wish to thank Prof. Paul Fitzgerald for providing clinical expertise, and all the patients who participated in the study.

## Patient Consent

All study procedures were approved by the Deakin University Human Research Ethics Committee (DUHREC 2021-136) and were conducted in accordance with the Declaration of Helsinki. All patients provided written informed consent to participate in the study.

