## Supplementary material for "Continuous theta-burst stimulation to a lesion network in cervical dystonia: a randomized, double-blind, sham-controlled, pilot trial": All supplementaries

**Corresponding authors:**

**SUPPLEMENTARY MATERIAL**

**Section 1: Supplementary Methods**

**Section 2: Supplementary Results**

**Figure S1.** CONSORT flow diagram of participant enrolment, randomization, and participation throughout the study.

**Figure S2.** Visualisation of Pearson's correlations between the raw change in MEP amplitude and raw change in TWSTRS total score, at each stimulator intensity across cTBS conditions.

**Figure S3.** Changes in right GPi functional connectivity following active cTBS.

**Table S1.** Participant guesses for which conditions they were allocated to in each block, and whether they guessed correctly.

**Table S2.** Uncorrected seed to whole brain fMRI analysis results.

### 1. Supplementary Methods

*1.1 Randomization*

Block order was randomised amongst patients using a complete randomisation protocol with the 'randomizr' package in R [1]. Randomization allocation was concealed via a password protected document that only the lead author (J.M-H.) had access to.

*1.2 Sample Size Calculations*

The projected sample size was guided by similar previous research which examined the effect of multi-day theta-burst stimulation to the cerebellum of CD patients [2, 3]. For a significant difference between pre and post-intervention TWSTRS scores, moderate effect sizes of $d$ = 0.31 [3] and $d$ = 0.42 [2] were reported. Working under the assumption that lesion network mapping increases efficacy for selection of treatment targets, a large effect size ($d$ = 0.8) was estimated for this study. Assuming 80% power (alpha level of 0.05), G*power (version 3.1) indicated a total sample size of 14 participants would be required. To ensure adequate power, and allowing for dropouts, we increased the sample to a target recruitment goal of 20 participants.

*1.3 Movement Kinematics and Proprioception Task Outcomes*

Triaxial gyroscope data recorded in degrees per second (º/s) were sampled at 60 Hz and processed in MATLAB R2025a (MathWorks Ltd, USA). Raw angular velocity signals (vx, vy, vz) were low-pass filtered using a zero-phase, 4th-order Butterworth filter with a cut-off frequency of 1Hz, to attenuate high-frequency noise while preserving voluntary movement-related components. Angular displacement vectors ($d_x$, $d_y$, $d_z$) were calculated by numerical integration of the filtered angular velocity signal using cumulative trapezoidal integration. The resulting signal was linearly de-trended to minimise drift, and zero-referenced to ensure all trials commenced from a common baseline.

Three kinematic outcome measures were extracted from each range of motion task:

1. *Range of motion* was calculated from the angular displacement signal as the difference between the maximum and minimum values over the duration of the trial.
2. *Peak angular velocity* was calculated from the absolute processed angular velocity signal using a peak detection algorithm. Peaks were identified using a prominence threshold set at 20% of the maximum signal amplitude, with a minimum inter-peak interval of 1s to avoid multiple detections within a single movement cycle. The mean of all detected peaks was calculated for each trial.
3. *Movement smoothness* was quantified using the Spectral Arc Length (SPARC) metric [4, 5]. For this analysis, raw angular velocity signals were low-pass filtered at 10Hz (rather than 1Hz) prior to computation to preserve higher-frequency involuntary components that might contribute to the smoothness estimate.

To characterise three-dimensional head orientation during the proprioception tasks, the triaxial angular displacement vectors ($d_x$, $d_y$, $d_z$) were used to compose the rotation matrix, ***R***, at all sampled timepoints (I) throughout the active and passive task conditions:

$$\boldsymbol{R_x}(t) = \begin{bmatrix} 1 & 0 & 0 \\ 0 & cos(d_x(t)) & -sin(d_x(t)) \\ 0 & sin(d_x(t)) & cos(d_x(t)) \end{bmatrix} \quad (1)$$

$$\boldsymbol{R_y}(t) = \begin{bmatrix} cos(d_y(t)) & -sin(d_y(t)) & 0 \\ sin(d_y(t)) & cos(d_y(t)) & 0 \\ 0 & 0 & 1 \end{bmatrix} \quad (2)$$

$$\boldsymbol{R_z}(t) = \begin{bmatrix} cos(d_z(t)) & 0 & sin(d_z(t)) \\ 0 & 1 & 0 \\ -sin(d_z(t)) & 0 & cos(d_z(t)) \end{bmatrix} \quad (3)$$

$$\boldsymbol{R}(t) = \boldsymbol{R}_x(t).\boldsymbol{R}_y(t).\boldsymbol{R}_z(t) \quad (4)$$

The matrix series ***R***(*t*) was then used to extract the unitary angular displacement value (θ), at each timepoint. The displacement angle (θ) is calculated as:

$$\theta(t) = arccos\left(\frac{tr(\boldsymbol{R}(t))-1}{2}\right) \quad (5)$$

For each trial, the target position was defined from the passive movement task. The time at which the examiner-imposed head position was reached was identified manually for each recording. The associated rotation matrix at this time point was then used to define the target orientation and to calculate the target angle (distance away from neutral).

During the subsequent active repositioning task, the participant's head orientation was similarly represented using rotation matrices. At each time point, the relative rotation between the active orientation and the target orientation was computed, and the angular difference extracted using the trace of the relative rotation matrix. This yielded a time series of angular error (in degrees) relative to the target position.

To ensure that accuracy reflected a sustained head position rather than transient passage through the target, a sliding window approach was applied. Angular error was evaluated over consecutive 1-second time windows, and for each window the maximum error was calculated. The minimum value across all 1-second windows was then taken as the smallest sustained positional error, representing the closest maintained alignment with the target for at least 1 second. This minimum angular error was then used as the outcome for analysis.

REML linear mixed effects models examined changes to minimum angular error, with timepoint and cTBS condition entered as fixed effects, and patient ID as the random intercept. Target angle of the passive movement task was included as a covariate to control for differences in passive head position amongst patients. Separate models were run for each of the movement direction axes (left, right, up, down).

*1.4 Magnetic Resonance Imaging*

All fMRI data preprocessing and analysis was performed using standardised pipelines with CONN (version 22; [6]) and SPM12 (https://fil.ion.ucl.ac.uk/spm/) toolboxes in MATLAB (version R2024b; Mathworks, USA). Functional and anatomical data were preprocessed using a modular preprocessing pipeline [7] including realignment with correction of susceptibility distortion interactions, slice timing correction, outlier detection, direct segmentation and MNI-space normalization, and smoothing. Functional data were realigned using SPM realign & unwarp procedure [8], where all scans were co-registered to a reference image (first scan of the first session) using a least squares approach and a 6 parameter (rigid body) transformation [9], and resampled using b-spline interpolation to correct for motion and magnetic susceptibility interactions. Temporal misalignment between different slices of the functional data (acquired in interleaved Siemens order) was corrected following SPM slice-timing correction (STC) procedure [10, 11], using sinc temporal interpolation to resample each slice BOLD timeseries to a common mid-acquisition time. Potential outlier scans were identified using ART [12] as acquisitions with framewise displacement above 0.9 mm or global BOLD signal changes above 5 standard deviations [13, 14], and a reference BOLD image was computed for each subject by averaging all scans excluding outliers. Functional and anatomical data were normalized into standard MNI space, segmented into grey matter, white matter, and CSF tissue classes, and resampled to 2 mm isotropic voxels following a direct normalization procedure [13, 15], using SPM unified segmentation and normalization algorithm [16, 17] with the default IXI-549 tissue probability map template. Lastly, functional data were smoothed using spatial convolution with a Gaussian kernel of 8 mm full width half maximum (FWHM).

Next, functional data were denoised using a standard denoising pipeline [18], including the regression of potential confounding effects characterized by white matter timeseries (5 CompCor noise components), CSF timeseries (5 CompCor noise components), motion parameters and their first order derivatives (12 factors; [19]), outlier scans (below 213 factors; [14]), session and task effects and their first order derivatives (8 factors), and linear trends (2 factors) within each functional run, followed by bandpass frequency filtering of the BOLD timeseries [20] between 0.008 Hz and 0.09 Hz. CompCor [21, 22] noise components within white matter and CSF were estimated by computing the average BOLD signal as well as the largest principal components orthogonal to the BOLD average, motion parameters, and outlier scans within each subject's eroded segmentation masks. From the number of noise terms included in this denoising strategy, the effective degrees of freedom of the BOLD signal after denoising were estimated to range from 136.2 to 276.8 (average 209.9) across all subjects [13].

Seed-based connectivity maps were estimated characterizing the patterns of functional connectivity from the six ROIs (bilateral and unilateral S1 cTBS stimulation sites: MNI x = ±45, y = -30, z = 58; bilateral and unilateral GPi: x = ±19.4, y = -10.1, z = -5.9 mm). Functional connectivity strength was represented by Fisher-transformed bivariate correlation coefficients from a weighted general linear model (weighted-GLM; [23]). Group-level analyses were performed using a General Linear Model (GLM). For each individual voxel a separate GLM was estimated, with first-level connectivity measures at this voxel as dependent variables (one independent sample per subject and one measurement per experimental condition), and groups or other subject-level identifiers as independent variables. Voxel-level hypotheses were evaluated using multivariate parametric statistics with random-effects across subjects and sample covariance estimation across multiple measurements. Inferences were performed at the level of individual clusters (groups of contiguous voxels). For clusters found in each fMRI analysis, threshold-free cluster enhancement (TFCE) was applied using 5000 permutations, with results considered significant with a family-wise error (FWE) corrected threshold of $p < .05$ [24, 25].

### 2. Supplementary Results

*2.1 Secondary Outcome: TWSTRS subscales*

A significant main effect of timepoint was found for the pain subscale ($F$(3, 82) = 3.46, $p$ =.020), but not for the severity ($p$ = .377) or disability subscales ($p$ = .233). *Post-hoc* pairwise comparisons demonstrated that this main effect was driven by significant reductions in TWSTRS pain scores from the Pre to the Post timepoint across the active and sham conditions ($b$ = -1.24; 95% CI: -0.26 – -2.21, $p_{corr}$ = .008; main manuscript Figure 2D).

*2.2 Secondary Outcome: Proprioception*

There were no significant timepoint × condition interaction effects for any of the movement axes in the proprioception task (left: $F$(63.13, 3) = 0.55, $p$ = .652; right: $F$(64.78, 3) = 0.21, $p$ = .889; up: $F$(62.95, 3) = 2.40, $p$ = .076; down: $F$(64.53, 3) = 1.52, $p$ = .218).

**Figure S1.** CONSORT flow diagram of participant enrolment, randomization, and participation throughout the study.

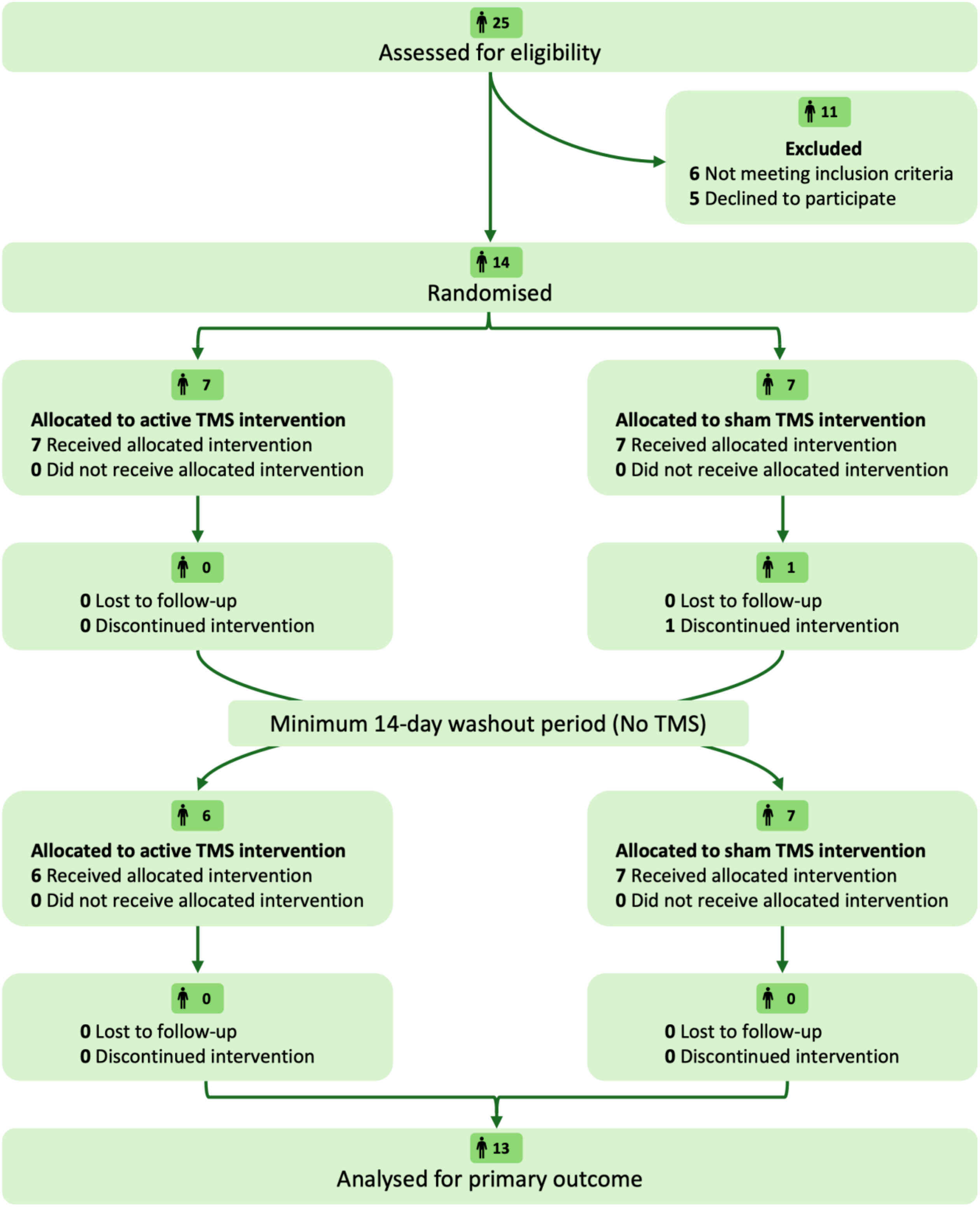

**Figure S2.** Visualisation of Pearson's correlations between the raw change in MEP amplitude and raw change in TWSTRS total score, at each stimulator intensity across cTBS conditions.

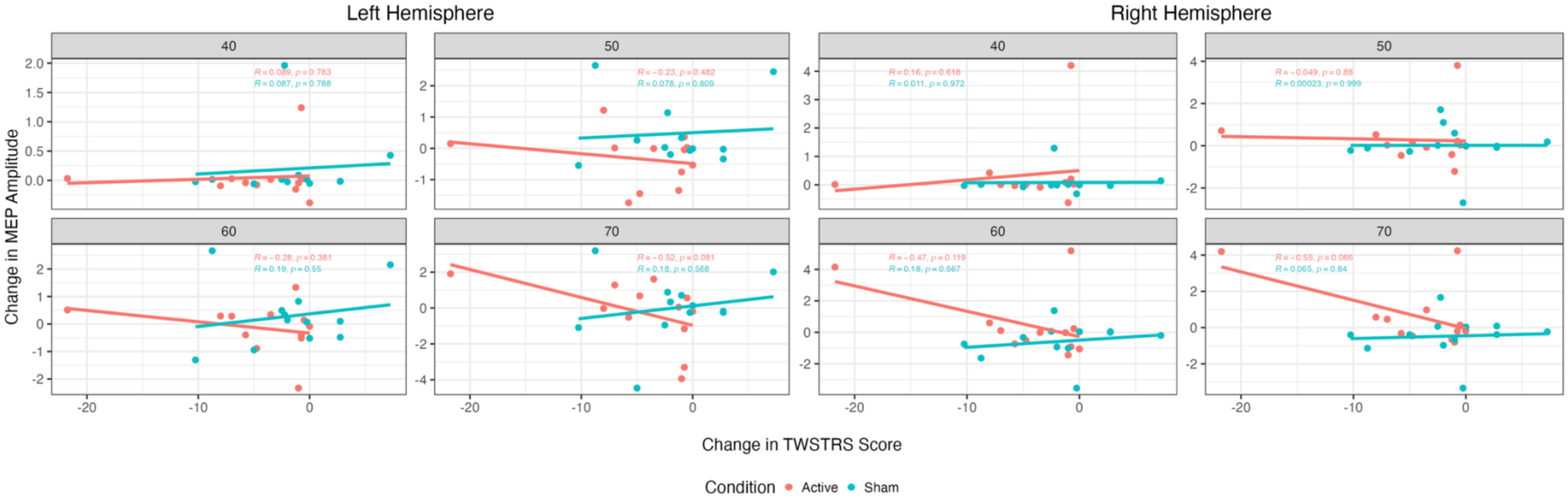

**Figure S3.** Changes in right GPi functional connectivity following active cTBS.

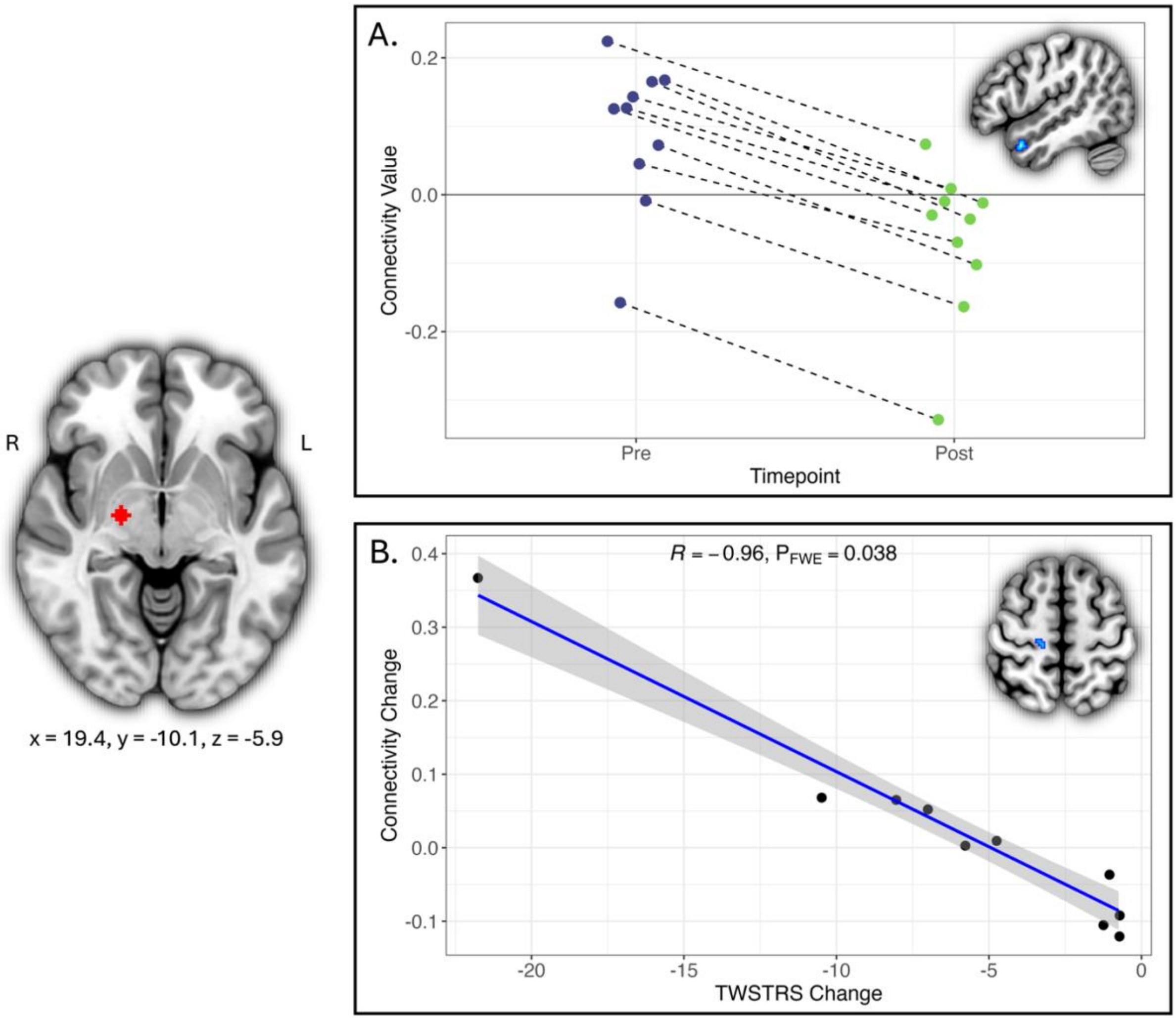


Figure A shows decreased positive connectivity from the right GPi to the right temporal pole (cluster centre of gravity: x = 50, y = 10, z = -28, cluster size: 33 voxels, whole brain $P_{FWE}$ = .012). Figure B shows the negative association between the right GPi–motor cortex connectivity and change in TWSTRS score (motor cortex centre of gravity: x = 14, y = -28, z = 58, 9 voxels, whole brain $P_{FWE}$ = .038).

**Table S1.** Participant guesses for which conditions they were allocated to in each block, and whether they guessed correctly.

| Subject ID | Correct guess? | Reason |
|---|---|---|
| 1 | Y | Initial funding could only support 2 scans per participant (additional funding was received prior to enrolment of other participants). Participant assumed we would schedule these around their active TMS block. |
| 2 | N | Had improvement in their leg dystonia in sham condition. |
| 3 | Y | Had finger twitch during active cTBS application, but not during sham. Also felt tremor improvement after active but not sham condition. |
| 4 | Y | Felt a slight intensity difference between the active and sham coils. |
| 5 | Y | Participant had a significant improvement in symptoms in the active condition but not in the sham. |
| 6 | Y | Participant felt a difference in the active and sham coils – the active coil felt more intense. |
| 7 | Y | Participant was unable to get an MRI scan before their sham block (due to scanner availability), and thus assumed we would schedule their MRI scans around their active block of TMS. |
| 8 | Y | Participant noted that in the sham block their symptoms felt like when their botox wears off. |
| 9 | Y | Participant noted that the active coil felt different (more intense) to the sham coil. |
| 10 | N | Participant did not guess the order of conditions correctly, and noted they had slight scalp discomfort for the day after their first session in both blocks. |
| 11 | Y | Participant felt a difference in the active and sham coils – the active coil felt more “intense”. Additionally, felt like in the sham stimulation block nothing changed for them at all, whereas across the active block they noticed changes to their symptoms. |
| 12 | N | Participant could not tell the difference between the sensations of the two coils. |
| 13 | Y | Participant felt a slight intensity difference between the coils. Additionally, some symptoms unrelated to dystonia changed in the active block. |

**Table S2.** Uncorrected seed to whole brain fMRI analysis results. Significant p-values are bolded.

| Contrast | ROI | Cluster Regions | Cluster Size (voxels) | Peak coordinate (MNI) | | | Peak uncorrected *p* |
|---|---|---|---|---|---|---|---|
| | | | | X | Y | Z | |
| **Active** | | | | | | | |
| | Bilateral GPi | Middle temporal gyrus, superior temporal gyrus, inferior temporal gyrus | 139 | 46 | 0 | -26 | < **.001** |
| | | Occipital pole, occipital fusiform gyrus, lateral occipital cortex | 132 | 26 | -96 | -16 | < **.001** |
| | R GPi | Temporal pole, middle temporal gyrus, superior temporal gyrus | 272 | 50 | 10 | -28 | < **.001** |
| | | Inferior frontal gyrus, precentral gyrus | 150 | -56 | 14 | 24 | < **.001** |
| | | Frontal pole, inferior frontal gyrus | 64 | -42 | 38 | 8 | < **.001** |
| *TWSTRS change* | | | | | | | |
| | R GPi | Precentral gyrus (M1), postcentral gyrus (S1) | 233 | 14 | -26 | 60 | < **.001** |
| **Sham** | | | | | | | |
| | Bilateral S1 | Frontal pole, superior frontal gyrus | 176 | 20 | 54 | 26 | < **.001** |
| | R GPi | Subcallosal cortex, cingulate/paracingulate gyrus | 74 | 6 | 26 | -6 | < **.001** |
| | R S1 | Frontal pole | 95 | 14 | 48 | 30 | < **.001** |
| *TWSTRS change* | | | | | | | |
| | L S1 | Occipital pole, lateral occipital cortex | 88 | 14 | -92 | 20 | < **.001** |
